# VIRAL LOAD SUPPRESSION AND ASSOCIATED FACTORS AMONG HIV-INFECTED PATIENTS ON SECOND-LINE ANTIRETROVIRAL THERAPY AT PUBLIC HEALTH FACILITIES OF WEST GUJI, GUJI AND BORENA ZONES, SOUTHERN ETHIOPIA: FACILITY BASED CROSS-SECTIONAL STUDY

**DOI:** 10.1101/2024.04.02.24305217

**Authors:** Digafe Hailu, Dube Jara, Alo Edin, Abdurazak Awol, Angefa Ayele, Yohannes Fekadu, Dereje Endale, Miesa Gelchu, Kebebew Lemma

## Abstract

**Background:** Ethiopia is one of the nation’s most severely impacted by HIV, with an estimated 700,000 people living with HIV/AIDS. Hence, many health facilities were providing second-line antiretroviral therapy, however little was known about viral load suppression among second-line users. This study aimed to assess the proportion of viral load suppression and associated factors among HIV-infected patients on second-line antiretroviral therapy at public health facilities of west Guji, Guji and Borena zones, Southern Ethiopia.

**Methods:** A facility-based cross-sectional study was conducted among 256 HIV-infected patients on second-line antiretroviral therapy from January 1, 2019, to December 30, 2022, by using census after obtaining ethical clearance from Bule Hora University ethical review committee. Data were extracted using a structured, pre-tested checklist, entered into the EPI data version 3.1.0, and exported to SPSS version 25 for analysis. The proportion of viral load suppression was determined. A binary logistic regression model was fitted to identify factors associated with viral load suppression. Statistical significance was declared at a 95% confidence interval (CI) with a P-value <0.05.

**Results:** This study revealed that the proportion of viral load suppression among HIV-infected patients on second-line antiretroviral therapy was 73.8% (95% CI, 68.0–79.1). Those who missed the second-line antiretroviral regimen [AOR = 0.315, 95% CI (0.162–0.612)], a baseline viral load count of <10,000 copies/mm3 [AOR = 2.291, 95% CI (1.216-4.316)], and a baseline body mass index of ≥18.5 kg/m2 [AOR = 2.438, 95% CI (1.098–5.414)] were significantly associated with viral load suppression.

**Conclusions:** The proportion of patients with viral load suppression fell below the WHO’s and national level. Viral load suppression was significantly influenced by missed second-line antiretroviral doses, a baseline viral load count of <10,000 copies/ml, and a baseline body mass index of ≥18.5 kg/m2. Hence interventions targeting counseling to patients that missed their antiretroviral therapy, keeping patient’s viral load to be less than 10,000 copies/ml through adequate adherence counseling among second-line antiretroviral therapy were recommended.

## Background

Antiretroviral regimens are recommended for use in order to achieve viral load suppression. The current recommendation for (antiretroviral) ARV regimens is to use three active drugs from two or more drug classes. The most significant indicator of initial and sustained response to ART is viral load (1). It is crucial to switch to an ARV regimen at the right time. Late switching can jeopardize the effectiveness of second-line therapy (2). Monitoring antiretroviral therapy (ART) patients is crucial to guarantee effective therapy, spot adherence issues, and decide whether switching ART regimens is necessary in the event of treatment failure. Since 2016, viral load testing has become the primary method for treatment monitoring, with 20 million viral load tests expected to be performed in low- and middle-income countries by 2020(3). Being on second-line ART, claiming to have poor adherence, and having a previously suppressed viral load were all independently linked to viral load suppression (4). Viral load monitoring among second-line ART can help reduce the burden on both patients and health care workers by reducing the need for frequent clinic visits (5).

A viral load that is undetectable and less than 50 copies/mL is known as viral load suppression (6). Globally, around 50 percent of HIV-infected patients were virally suppressed (7). Study findings of cohort study in Africa showed that among PLWHA with ART experience and viral suppression, those who moved to tenofovir/lamivudine/dolutegravir (TLD) maintained viral suppression better than those who persisted on other regimens (8). The HIV program in Kenya reported that second-line ART users had a viral load suppression rate of 90% (9). Findings revealed that among 46.8% of HIV patients on second-line ART, 88.2% achieved viral load suppression (4). Ethiopia’s findings revealed that second-line ART had a viral load suppression rate of 82.39% (10). There is a paucity of data in Ethiopia and in many resource-limited settings for viral load suppression among HIV-infected patients who received second-line ART. Evidence on the proportion of viral load suppression will provide input to policy and decision makers, program planners, and implementers (both governmental and nongovernmental organizations) for monitoring and evaluation activities (to assess the implementation of national ART guidelines and proper delivery of second-line ART in improving patients’ quality of life), to create awareness of the risks, and to take appropriate actions on the factors that promote viral load suppression. This study will also increase the public health body of knowledge and help promote researchers in this area. Moreover, the findings will be used as an additional source of information for further research on related subject matter. In Ethiopia, second-line ART is being implemented in many health facilities; however, there is a single study conducted among second-line ART in the northern part of Ethiopia. This study was limited in scope to further generalize the findings. Therefore, the study is important to fill these research gaps by covering a large study area. Therefore, the aim of this study is to assess the proportion of viral load suppression and associated factors among HIV-infected patients on second-line ART at public health facilities of west Guji, Guji and Borena zones, Southern Ethiopia, from January 1, 2019, to December 30, 2022.

## Methods

### Study Area, Study Design and Study Period

A facility-based cross-sectional study design was conducted in west Guji, Guji and Borena zones, southern Ethiopia from January 01, 2019 to December 30, 2022. The actual data collection period was from June 20, 2023 to July 10, 2023. West Guji is located 467km, Guji is located 599km and Borena is located 566km away from a capital city of Ethiopia (Addis Ababa) with a total population of 3,776,556. West Guji has 4 hospitals and 42 health centers from these two hospitals and one health center was centers for second-line ART service provision. Guji zone has 4 hospitals and 62 health centers, from these 2 hospitals and 2 health centers were center for second-line ART service provision. Borena zone has 5 hospitals and 45 health centers, from these two hospitals and one health center were providing second-line ART.

### Populations

The source populations of this study was all HIV-infected patients on second-line antiretroviral therapy at public health facilities of west Guji, Guji and Borena zones, southern Ethiopia. The study populations was all HIV-infected patients on second-line antiretroviral therapy at public health facilities of west Guji, Guji and Borena zones, Southern Ethiopia from January 1, 2019 to December 30, 2022. All HIV-infected patients’ on second-line antiretroviral therapy for more than 6 months of duration at public health facilities of west Guji, Guji and Borena zones, Southern Ethiopia from January 1, 2019 to December 30, 2022 were included in the study whereas those HIV-infected patients, on second-line ART with an unknown viral load test result after the initiation of second-line ART, lost to follow up and those with incomplete charts were excluded from the study.

### Sample size Determination

The sample size is calculated by using Epi Info version 7.0 software sample size calculation program for cross sectional study. Considering the following assumptions: 5% level of significance, power of 80% and the different exposure variables like Baseline Viral load count <10,000coppies, Isoniazid preventive therapy (yes), and Adherence to ART (good) from previous study. Finally, by adding 10% non-response rate the total sample size was 245.

### Sampling method and procedure

Of the ten health facilities expected to provide second-line ART service, eight of them were providing second-line ART in west Guji, Guji, and Borena zones, Southern Ethiopia. These were Bule Hora University Specialized Hospital, Yabelo General Hospital, Moyale Primary Hospital, Moyale Health Center, Negele General Hospital, Adola General Hospital, Adola Health Center, and Shakiso Health Center. From January 1, 2019 through December 30, 2022, a total of 256 HIV-infected patients received second-line antiretroviral therapy. Since the study populations that fulfill the inclusion criteria were below the predicted proportion, all the study populations in the chosen study area were taken into consideration as a sample population. A study unit was selected using census following thorough chart review.

### Study variables

**The dependent variable** of this study was **Viral Load Suppression**.

### The independent variables were

**Socio-demographic Factors: *-***Age, Sex, Marital Status, Educational Status, Residence, Religion, and Types of Health Institutions.

### Baseline clinical and laboratory-related factors

Baseline CD4, Baseline viral load, WHO clinical stages, Baseline weight, Baseline BMI, Functional status, TB at baseline, Other OI at baseline, Admission history.

### Treatment-related factors

Cotrimoxazole preventive therapy, Isoniazid preventive therapy, antiretroviral dose per day, ART regimen, Duration on ART.

### Behavioral-related factors

Adherence level to antiretroviral therapy, Disclosure status, Missing ARV dose.

### Operational definitions

**Baseline viral load:** Viral load result at the start of second-line antiretroviral therapy

**Baseline CD4:** CD4 count at the start of the second line ART

**Baseline clinical stage:** WHO stage at the start of the second line ART **Viral load suppression:** viral load test results of less than 50 copies/mm3 **Baseline Functional Status** (11):

**Working:** able to perform usual work inside or outside the home **Ambulatory:** able to perform activities of daily living; not able to work **Bedridden:** not able to perform activities of daily living

### ART adherence level(12)

**Good adherence:** average adherence ≥ 95% (patient missing ≤2 doses from 30 doses or ≤3 doses out of 60 doses).

**Fair adherence:** average adherence of 85–94% (patient missing 2-4 doses out of 30 doses or 4– 9 doses from 60 doses).

**Poor adherence:** average adherence < 85% (patient missing ≥5 doses from 30 doses or ≥10 doses from 60 doses).

**Missed second-line antiretroviral therapy:** when the patient had poor adherence to ARV regimens.

### Data collection tools and procedures

Using a structured checklist, data gatherers retrieved secondary information from the patient’s chart, viral load registration logbook, and laboratory request sheets. As a result, all patient charts that contained comprehensive data about those who received second-line antiretroviral therapy were examined. When incomplete data was discovered, the data gatherers attempted to obtain the information from several data sources (such as the client’s chart and follow-up form). Additionally, the data that was most current to the start date of second-line antiretroviral medication was deemed baseline data when clinical parameters and laboratory results (CD4 count and WHO clinical stages) were not found at the start of second-line antiretroviral therapy. Eight nurses with a Bachelor of Science in nursing engaged in the data extraction process as data collectors, and four ART data managers served as supervisors for the three zones.

### Data quality assurance

Data quality was assured through the design of a proper data extraction tool. Both data collectors and supervisors received training for one day on the objectives of the study and how to extract data from patient charts, viral load registration logbooks, and laboratory request sheets. The pretest was done by taking 5% of the overall sample at Dilla University General Hospital. During the data collection, a supervisor was assigned to make sure that there was no missed data. The study’s investigator was in charge of the entire data extraction process. The reliability (internal consistency) of the tools was measured using Cronbach alpha as the variables were adapted from different literature, and the result was 0.751, indicating that the variables were internally consistent. To check for any errors, missing values, and outliers, the data was cleaned in the end, and analysis was done.

### Data processing and analysis

The accuracy and completeness of the data were verified before being entered into Epi Data version 3.1.0 and exported to SPSS version 25 for analysis. The characteristics of the study participants were described using descriptive statistics such as frequencies and percentages for categorical variables and mean and standard deviation for continuous variables. Binary logistic regression analysis was used to identify the association between dependent and independent variables. All independent variables that show association at bivariable logistic regression with a P-value of <0.25 were considered for multivariable logistic regression analysis to identify factors independently associated with viral load suppression after adjusting for possible confounders. The variance inflation factor (VIF), which was used to test for multi-collinearity amongst independent variables, is less than five for all variables used in the analysis. The model passed the Hosmer-Lemeshow goodness-of-fit test with a score of 0.619. Finally, the AOR determined the level of significance with a 95% confidence interval and a P-value of< 0.05.

## Results

### Socio-demographic characteristics of the study participants

A total of 256 charts of HIV-infected patients on second-line antiretroviral therapy were reviewed with a response rate of 100%. The majority, 136 (or 53.1%), were married, and 139 (54.3%) of them were women. In terms of educational attainment, the majority of participants 128 (50%) had at least a primary education, whereas 161 (62.9%) of the study’s participants reside in an urban region. Nearly half of the research participants 119 or 46.5% received second-line ART in the general hospital. Overall, 162 (63.3%) of the participants belong to the age group of ≥ 36 years (Table 1).

**Table 1:** Socio-demographic characteristics of HIV-infected patients on second-line antiretroviral therapy at public health facilities of west Guji, Guji and Borena zones, Southern Ethiopia, in 2024 (n = 256)

| Variables | Categories | Frequency (n) | Percent (%) |
| --- | --- | --- | --- |
| **Age in years** | 6-15 | 15 | 5.9 |
|  | 16-25 | 32 | 12.5 |
|  | 26-35 | 47 | 18.4 |
| | $\geq 36$ | 162 | 63.2 |
| **Sex** | Male | 117 | 45.7 |
|  | Female | 139 | 54.3 |
| **Marital status** | Single | 56 | 21.9 |
|  | Married | 136 | 53.1 |
|  | Divorced | 40 | 15.6 |
|  | Widowed | 24 | 9.4 |
| **Educational status** | Cannot read and write | 79 | 30.9 |
|  | Primary education | 128 | 50.0 |
|  | Secondary education and above | 49 | 19.1 |
| **Residence** | Urban | 161 | 62.9 |
|  | Rural | 95 | 37.1 |
| **Religion** | Orthodox | 97 | 37.9 |
|  | Muslim | 55 | 21.5 |
|  | Protestant | 91 | 35.5 |
|  | Others | 13 | 5.1 |
| **Types of health institutions** | Health Center | 44 | 17.2 |
|  | Primary Hospital | 12 | 4.7 |
|  | General Hospital | 119 | 46.5 |
|  | Specialized Hospital | 81 | 31.6 |

### Baseline clinical and laboratory characteristics of the study participants

More than half 177 (69.1%) of study participants had a baseline CD4 count of >200 cells/mm3, whereas 129 (50.4%) participants had a baseline viral load count of >=10,000 copies. The majority of the respondents, 167 (65.2%), were stage I RVI, and 200 (78.1%) were working. With regard to baseline weight, 149 (58.2%) of the study participants were >50 kg (Table 2).

**Table 2:** Baseline clinical and laboratory-related characteristics of HIV-infected patients on second-line ART at public health facilities of west Guji, Guji and Borena zones, SouthernEthiopia, in 2024 (n = 256)

| **Variables** | **categories** | **Frequency (n)** | **Percent (%)** |
| --- | --- | --- | --- |
| **CD4 cell count** | ≤200cells/mm<sup>3</sup> | 79 | 30.9 |
|  | >200cells/mm<sup>3</sup> | 177 | 69.1 |
| **Viral load count** | <10,000 copies/mm3 | 127 | 49.6 |
|  | ≥10,000 copies/mm3 | 129 | 50.4 |
| **WHO RVI stage** | Stage I | 167 | 65.2 |
|  | Stage II | 19 | 7.4 |
|  | Stage III | 57 | 22.3 |
|  | Stage IV | 13 | 5.1 |
| **Weight in kg** | ≤20kg | 14 | 5.5 |
|  | 21-50kg | 93 | 36.3 |
|  | >50kg | 149 | 58.2 |
| **BMI in kg/m2** | <18.5kg/m2 | 93 | 36.3 |
|  | ≥18.5kg/m2 | 163 | 63.7 |
| **Functional status** | Working | 200 | 78.1 |
|  | Ambulatory | 38 | 14.8 |
|  | Bedridden | 18 | 7.1 |
| **Tuberculosis(TB)** | No | 231 | 90.2 |
|  | Yes | 25 | 9.8 |
| **Other OI** | No | 232 | 90.6 |
|  | Yes | 24 | 9.4 |
| **Admission history** | No | 220 | 85.9 |
|  | Yes | 36 | 14.1 |

### Treatment-related characteristics of the study participants

The majority, 218 (85.2%) of study participants, used CPT, and 202 (78.9%) of them used IPT. Nearly half of respondents, 116 (45.3%), were on AZT-3TC-ATV/r. Almost all 230 (89.8%) of the patients took their second-line ART twice daily (Table 3).

**Table 3:** Treatment-related characteristics of HIV-infected patients on second-line ART at public health facilities of west Guji, Guji and Borena zones, Southern Ethiopia, in 2024 (n = 256)

| Variables | Categories | Frequency (n) | Percent (%) |
| --- | --- | --- | --- |
| **CPT** | No | 38 | 14.8 |
|  | Yes | 218 | 85.2 |
| **IPT** | No | 54 | 21.1 |
|  | Yes | 202 | 78.9 |
| **Second-line ART doses per day** | Once | 26 | 10.2 |
|  | Twice | 230 | 89.8 |
| **Second-line ART** | AZT-3TC-LPV/r | 15 | 5.9 |
|  | AZT-3TC-ATV/r | 116 | 45.3 |
|  | TDF-3TC-LPV/r | 24 | 9.4 |
|  | TDF-3TC-ATV/r | 75 | 29.3 |
|  | ABC-3TC-LPV/r | 14 | 5.5 |
|  | ABC-3TC-DTG(5M) | 12 | 4.7 |
| **Duration on second-line ART** | ≤24 months | 59 | 23.0 |
|  | >24 months | 197 | 77.0 |

**CPT:** Cotrimoxazole preventive therapy; **IPT:** Isoniazid preventive therapy.

### Behavioral-related characteristics of the study participants

The majority of research participants, 181 (70.7%), had good adherence to second-line antiretroviral medication. Seventy-seven percent of research participants told their families they were HIV positive. Two-thirds (75.0%) of the subjects did not miss their second-line antiretroviral therapy (Table 4).

**Table 4:**
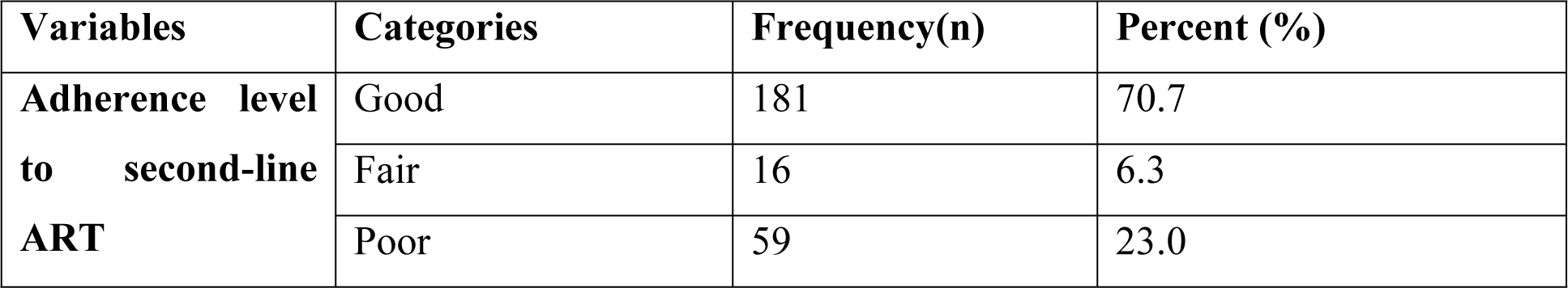
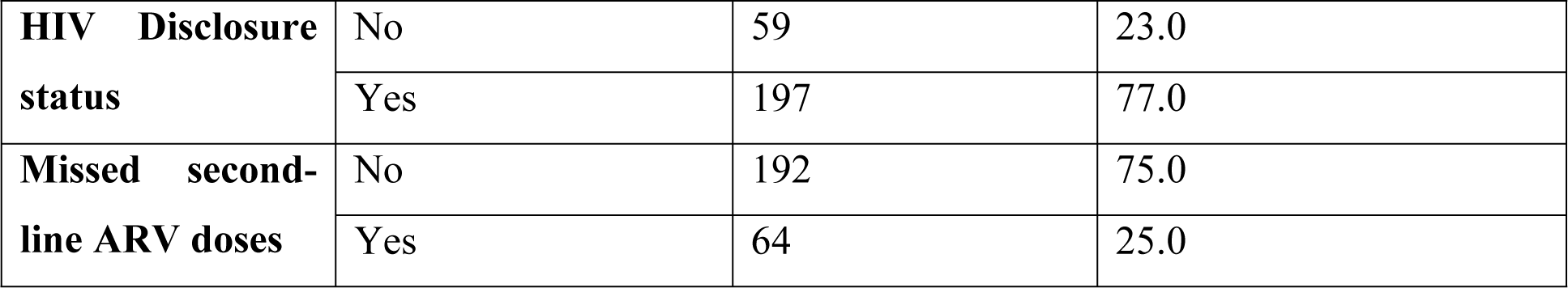
Behavioral-related characteristics of HIV-infected patients on second-line ART at public health facilities of west Guji, Guji and Borena zones, Southern Ethiopia, in 2024 (n = 256)

### Proportion of viral load suppression

The proportion of viral load suppression among HIV-infected patients who received second-line antiretroviral therapy was 73.8% [95% CI (68.0–79.1)], which is nearly two-thirds of the study populations (Figure 1).

**Figure 1:**
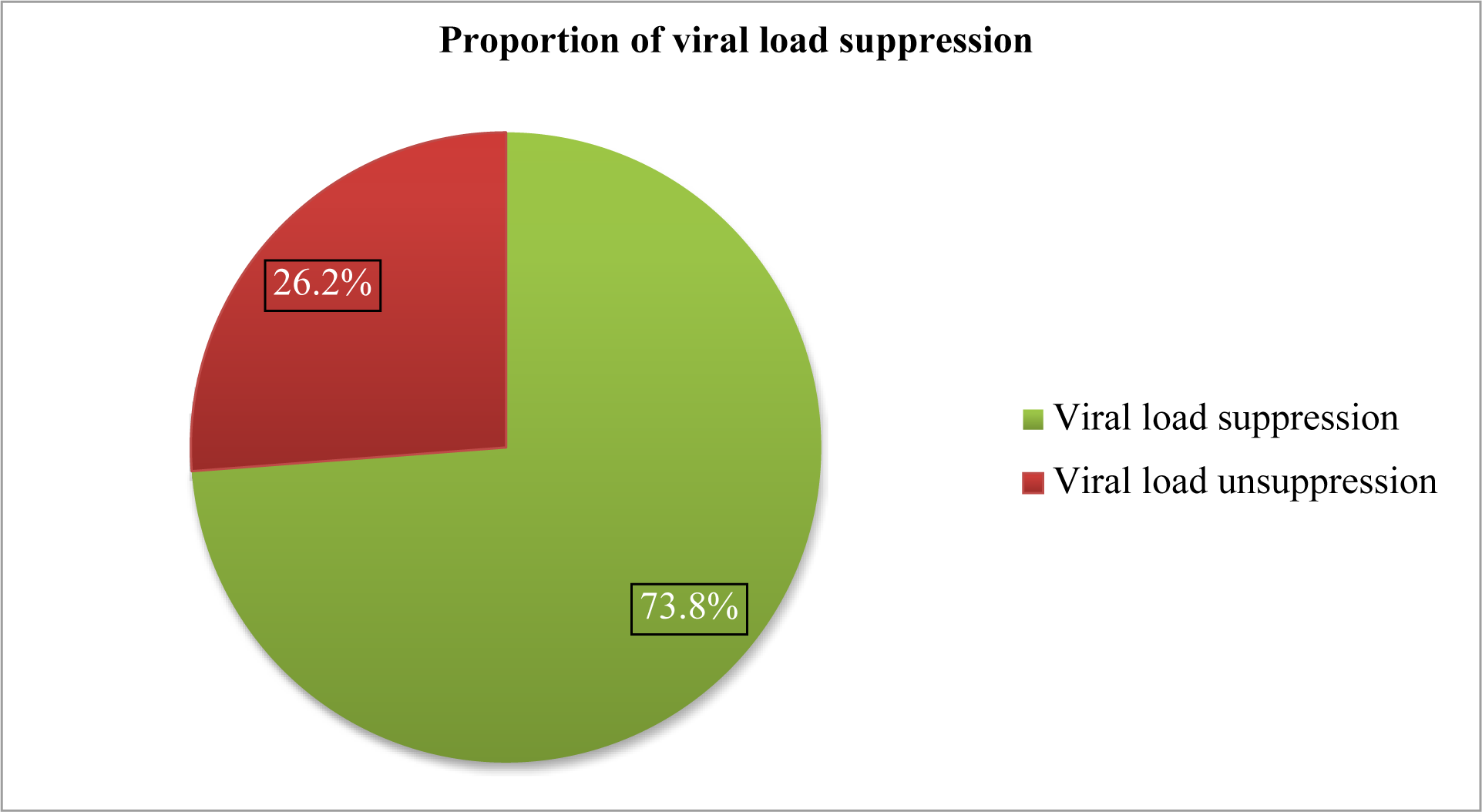
Proportion of viral load suppression among HIV-infected patients on second-line antiretroviral therapy at public health facilities of west Guji, Guji and Borena zones, Southern Ethiopia, in 2024 (n = 256)

### Factors associated with viral load suppression

A bi-variable logistic regression was carried out to identify potential variables for a multivariable logistic regression. Age, marital status, residence, missed second-line antiretroviral doses, baseline viral load count, baseline weight, baseline BMI, and duration of second-line antiretroviral therapy were found to have a significant association with the outcome variable at a p-value less than 0.25 and were considered candidate variables for multivariable logistic regression. Accordingly, these variables were selected as candidates for the initial multivariable logistic regression, and three variables: - missed second-line antiretroviral doses, a baseline viral load count of <10,000 copies/mm3, and a baseline body mass index of ≥18.5 kg/m2 were found to have a statistically significant association with the outcome variable at a p-value of less than 0.05 in the multivariable analysis. Those participants who missed their second-line antiviral medications were 68.5% less likely to experience viral load suppression than those who did not miss their medication [AOR = 0.315, 95% CI (0.162–0.612)]. Study participants with a baseline viral load count of <10,000 copies/mm3 had a 2 times higher chance of achieving viral load suppression compared to those with a baseline viral load count of ≥10,000 copies/mm3 [AOR = 2.291, 95% CI (1.216–4.316)]. Similarly, participants with a baseline body mass index of ≥18.5 kg/m2 had about a two-fold increased chance of achieving viral load suppression than those with a baseline body mass index of <18.5 kg/m2 [AOR = 2.438, 95% CI (1.098–5.414)] (Table 5).

**Table 5:** Bi-variable and multivariable analysis of factors associated with viral load suppression among HIV-infected patients on second-line antiretroviral therapy between January 1, 2019 and December 30, 2022, at public health facilities of west Guji, Guji and Borena zones, Southern Ethiopia, in 2024 (n = 256)

| Variables | Categories | Viral load status |  | COR(95% CI) | AOR(95% CI) | P-value |
| --- | --- | --- | --- | --- | --- | --- |
|  |  | Suppressed(n) | Unsuppressed(n) |  |  |  |
| Age in years | 6-15 | 9 | 6 | 0.398(0.133-1.197) | 0.747(0.133-4.178) | 0.740 |
|  | 16-25 | 18 | 14 | 0.342(0.154-0.756) | 0.384(0.114-1.310) | 0.127 |
|  | 26-35 | 34 | 13 | 0.695(0.331-1.460) | 0.679(0.300-1.536) | 0.353 |
|  | ≥36 | 128 | 34 | **1** | **1** |  |
| **Marital status** | Single | 35 | 21 | 0.152(0.032-0.711) | 0.354(0.052-2.394) | 0.287 |
|  | Married | 103 | 33 | 0.284(0.063-1.271) | 0.251(0.049-1.288) | 0.097 |
|  | Divorced | 29 | 11 | 0.240(0.048-1.193) | 0.207(0.036-1.188) | 0.077 |
|  | Widowed | 22 | 2 | **1** | **1** |  |
| **Residence** | Urban | 123 | 38 | 1.422(0.806-2.511) | 1.439(0.756-2.741) | 0.268 |
|  | Rural | 66 | 29 | **1** | **1** |  |
| **Missed second-line ART** | No | 153 | 39 | **1** | **1** |  |
|  | Yes | 36 | 28 | 0.328(0.179-0.601) | **0.315(0.162-0.612)** | **0.001** |
| **Duration on second-line ART** | ≤24months | 38 | 21 | 0.551(0.295-1.032) | 0.515(0.257-1.031) | 0.061 |
|  | >24months | 151 | 46 | **1** | **1** |  |
| **Baseline weight in kg** | ≤20kg | 6 | 8 | 0.222(0.072-0.683) | 0.562(0.104-3.034) | 0.503 |
|  | 21-50kg | 68 | 25 | 0.804(0.443-1.461) | 1.656(0.714-3.837) | 0.240 |
|  | >50kg | 115 | 34 | **1** | **1** |  |
| **Baseline viral load count** | <10,000 copies | 103 | 24 | 2.146(1.207-3.816) | **2.291(1.216-4.316)** | **0.010** |
|  | ≥10,000 copies | 86 | 43 | **1** | **1** |  |
| **Baseline body mass index** | <18.5kg/m<sup>2</sup> | 60 | 33 | **1** | **1** |  |
|  | ≥18.5kg/m<sup>2</sup> | 129 | 34 | 2.087(1.182-3.684) | **2.438(1.098-5.414)** | **0.029** |

The Arabic number (**1**) represents “reference”

## Discussions

This study was conducted to determine the proportion of viral load suppression and associated factors among HIV-infected patients on second-line antiretroviral therapy. The findings of the study suggest that the overall viral load suppression among HIV-infected patients on second-line antiretroviral therapy was 73.8% (95% CI: 68.0–79.1). This finding is consistent with studies conducted in the Ehlanzeni district of South Africa (74%), Uganda (75.8%), Kumasi, Ghana (76.1%), and the East Shewa zone of Oromia, Ethiopia (72%)(13–16). However, this finding is lower than those of studies conducted in the USA (86%)(17), Buenos Aires, Argentina (87.3%)(18), Cameroon (88.2% and 94.55%)(19, 20), Uganda (81%, 85%, & 85.7%)(21–23), Bamako, Mali (100%)(24), Mazowe, Zimbabwe (94.4%)(25), Tanzania (91.5%)(26), South Africa (77% and 77.2%)(27, 28), Kenya (80% and 90%) (29, 30), Ethiopia (82% and 82.39%) (10, 31). The ambitious 95-95-95 approach put forth by USAID, WHO, and UNAIDS to achieve 95% viral load suppression by 2030 is too far from the current results (5, 32). This discrepancy could be attributed to a variety of factors, including differences in the resources available to facilities and the study’s exclusive focus on second-line antiretroviral therapy.

In contrast to the above, the current finding is higher than the findings reported from the studies done in the United States (30.51%), Zimbabwe (30%), Johannesburg, South Africa (56.5%), Kampala, Uganda (66.4%), Gomba district, rural Uganda (10%), Haut-Katanga and Kinshasa Provinces of the Democratic Republic of the Congo (56.4%), Wollo, Northeast Ethiopia (66.4%), Bahir Dar, Ethiopia (57.1%), and the Public Hospitals of Hawasa City Administration, Ethiopia (40.9%)(11, 32–39). This discrepancy could be due to the fact that different donors are currently working in Ethiopia in collaboration with government to achieve targets in viral load suppression rate for 2030.

The current study found that among HIV-infected patients on second-line antiretroviral therapy and those who missed second-line antiretroviral regimens, a baseline viral load count of <10,000 copies/mm3 and a baseline body mass index of ≥ 18.5 kg/m2 were statistically significant factors associated with viral load suppression. The findings of the study showed that HIV-infected individuals who missed second-line antiretroviral therapy were less likely to experience viral load suppression than HIV-infected individuals who did not miss their second-line antiretroviral therapy. This may be due to the fact that skipping antiretroviral doses might cause HIV to change its form, impairing the effectiveness of therapy by making drugs less effective and increasing the risk of developing drug resistance. When the ARV regimen is missed, HIV has a chance to grow quickly and impair the immune system, which keeps the viral load high(40). As a result, an HIV-infected patient who follows the recommended HIV treatment regimen can stay healthy and prevent HIV transmission to sex partners(41). This result is consistent with studies done in India, San Francisco, resource-limited settings, Uganda, Malawi, Zimbabwe, and South Africa (42–47).

This study demonstrates that baseline viral load count is a significant factor that influences the viral load suppression process. HIV-infected patients on second-line antiretroviral therapy and had a baseline viral load count of <10,000 copies/mm3 show a 2-fold increased chance of achieving viral load suppression as compared to those who had a baseline viral load count of ≥10,000 copies/mm3.This disparity between the groups is brought about by the fact that when the viral load is low at baseline, the virus is likely not actively reproducing as quickly, immune system damage may be reduced, and HIV consequences may be less common, all of which contribute to a lower HIV reservoir in the blood(48).This finding is in line with research from Taiwan, Uganda, Wollo, Bahir Dar, and Arba Minch, Ethiopia(11, 35, 37, 49–52).

Viral load suppression was significantly influenced by body mass index (BMI). Patients on second-line antiretroviral medication and whose baseline BMI was ≥18.5 kg/m2 had a higher chance of experiencing viral load suppression compared to patients whose baseline BMI was <18.5 kg/m2.This can be explained by the fact that when a person’s BMI is low or less than 18.5 kg/m2, as compared to a normal BMI, they are more susceptible to malnutrition and compromised immune function, which raises their risk of contracting a number of diseases. As a result, viral load levels will increase when the immune system is weak or sick, limiting the likelihood of viral load suppression(53). Patients with a low BMI should therefore get appropriate dietary therapy and counseling in accordance with established treatment protocols. This finding is in line with the research conducted in South Wollo, Ethiopia(49).

## Limitations of the study

Secondary data makes it challenging to locate patient information quickly and to get adequate variables from the charts.

Due to the small number of people taking second-line antiretroviral therapy, a wider range of age groups (including both children and adults) was chosen to increase the number of participants.

## Conclusions

The proportion of viral load suppression was less than what the WHO and UNAIDS strategy had suggested for 2030. A missed second-line antiretroviral regimen, a baseline viral load count < 10,000 copies/mm3, and a baseline body mass index ≥ 18.5 kg/m2 were found to have a statistically significant and independent association with viral load suppression. Hence interventions targeting providing counseling to patients that missed their therapy, keeping patient’s viral load to be less than 10,000 copies/ml through adequate adherence counseling among second-line antiretroviral therapy were recommended.

## Ethical approval

To grant access to patient records, Bule Hora University Specialized Hospital, Yabelo General Hospital, Moyale Primary Hospital, Moyale Health Center, Negele General Hospital, Adola General Hospital, Adola Health Center, and Shakiso Health Center received ethical clearance from ethical review committee of Bule Hora University, the Institute of Health with reference number of BHU/IOH/SPH/0039/2015.

## Consent of participants

Data were taken from the patient’s chart, laboratory request sheets, and high viral load registration logbook after obtaining consent from ART focal person of each health facility. As the data was obtained from patient records and its source was secondary data, obtaining consent from study participants were not possible. Therefore, data confidentiality was maintained throughout the study, and in accordance with the Helsinki Declaration, the data will not be utilized for any other purposes.

## Consent for publication

Not applicable.

## Availability of data and materials

All relevant data are available from the corresponding author upon reasonable request

## Competing interests

The authors declare that they have no competing interests

## Funding

Bule Hora University

## Authors’ Contribution

DH: Designed and participated in data collection, conducted the data analysis and interpretation, developed the first draft and revised subsequent drafts. DJ: Advised on the conception of study area, data analysis and interpretation reviewed and commented on successive drafts. AE: Advised on the data analysis and interpretation and commented on successive drafts and reviewed and edited. AbA: Advised on the conception of study area, data analysis and interpretation reviewed and commented on successive drafts. AA: Advised on the data analysis and interpretation and commented on successive drafts and reviewed. YF: Participated in data collection and developed the first draft and revised subsequent drafts. DE: Advised on the conception of study area, data analysis and interpretation. Reviewed, edited and commented on successive drafts. MG: Advised on the successive drafts reviewed and participated in analysis. KL: Advised on the conception of study area, data analysis and interpretation reviewed and commented on successive drafts. All authors reviewed and approved the final manuscript.

## Data Availability

All relevant data are available from the corresponding author upon reasonable request

## Acknowledgments

The authors acknowledge Bule Hora University Institute of Health, School of Public Health, for the overall facilitation. We would like to thank the data collectors and supervisors for their contribution to materializing data collection activities and sincerely thanking Meriyan Abdulkadir, Kasu Dade, and Firomsa Abera for their assistance in sharing data and up-to-date information about second-line antiretroviral therapy.

## Abbreviations

AIDS: Acquired Immunodeficiency Syndrome
ART: Antiretroviral Therapy
ARV: Antiretroviral
BMI: Body Mass Index
CPT: Co-trimoxazole Preventive Therapy
DTG: Dolutegravir
DVS: Durable Viral Suppression
FDC: Fixed Dose Combination
HAART: Highly Active Anti-retroviral Therapy
HIV: Human Immunodeficiency Virus
HTS: HIV Testing Service
IL: Interleukin
IPT: Isoniazid Preventive Therapy
MDGs: Millennium Development Goals
NRTI: Non Reverse Transcriptase Inhibitor
OI: Opportunistic Infection
PI: Protease Inhibitor
PLWHA: People Living With HIV/AIDS
RNA: Ribonucleic Acid
RVI: Retroviral Infection
TB: Tuberculosis
tVR: transient Viral Rebound
VIF: Variance Inflation Factor
VLS: Viral Load Suppression

## Notes

### Competing Interest Statement

The authors have declared no competing interest.

### Funding Statement

Funding was secured from Bule Hora University, where I earned my MPH. Bule Hora University approved title for use and does not have any role in design of study. collection, analysis, interpretation of data, and writing of the manuscript.

### Author Declarations

To grant access to patient records, Bule Hora University Specialized Hospital, Yabelo General Hospital, Moyale Primary Hospital, Moyale Health Center, Negele General Hospital, Adola General Hospital, Adola Health Center, and Shakiso Health Center received ethical clearance from ethical review committee of Bule Hora University, the Institute of Health with reference number of BHU/IOH/SPH/0039/2015. Data were taken from the patient's chart, laboratory request sheets, and high viral load registration logbook after obtaining consent from ART focal person of each health facility. As the data was obtained from patient records and its source was secondary data, obtaining consent from study participants were not possible. Therefore, data confidentiality was maintained throughout the study, and in accordance with the Helsinki Declaration, the data will not be utilized for any other purposes.

